# COMORBIDITY OF PULMONARY TUBERCULOSIS AND BACTERIAL PNEUMONIA IN PATIENTS WITH LATE STAGES OF HIV-INFECTION WITH IMMUNODEFICIENCY

**DOI:** 10.1101/2022.12.07.22283221

**Authors:** A.V. Mishina, V.Yu. Mishin, I. V. Shashenkov

## Abstract

**Purpose:** To study specifics of diagnostics and clinics of comorbidity of pulmonary tuberculosis and bacterial pneumonia in patients with HIV-infection with immunodeficiency.

**Materials and methods:** Ninety-three first-time diagnosed patients with pulmonary tuberculosis and 4B stage of HIV-infection in the advanced phase in the absence of antiretroviral therapy were examined. The patients were divided into 3 groups. The 1^st^ group included 31 patient with pulmonary tuberculosis and pneumonia associated with *Streptococcus pneumoniae*, the 2^nd^ group included 31 patient with pulmonary tuberculosis and pneumonia associated with *Staphylococcus aureus*. The 3^rd^ group included 31 patient without bacterial pneumonia selected by a copy-pair principle. Statistical treatment of the data was performed using Microsoft Office Excel 2019 with calculation of the mean parameter in the group and the standard error of the mean confidence interval (CI).

**Results:** Comorbidity of pulmonary tuberculosis and pneumonia associated with *S. pneumoniae* or *S. aureus* in patients with 4B stage of HIV-infection with immunodeficiency in the advance phase with absence of antiretroviral treatment is characterized with generalization of tuberculosis and development of opportunistic infections of the lungs with severe clinical picture, high level of drug resistance of *M. tuberculosis* and the agents of bacterial pneumonia. At computed tomography of the chest a focal dissemination is revealed in the lungs as well as an intrathoracic lymphadenopathy and changes of the lung pattern, which almost does not differ in patients with bacterial pneumonia.

**Conclusion:** Clinical signs and X-ray changes in combination of pulmonary tuberculosis and pneumonia associated with *S. pneumoniae* or *S. aureus* and pulmonary tuberculosis with bacterial pneumonia at the late stages of HIV-infection with immunodeficiency have the same type of character that can be diagnosed only with special microbiological viral and molecular genetic studies of abnormal material from the respiratory system and other organs with obligatory determination of drug resistance to the antituberculosis drug products and the antibacterial agents of wide spectrum.

## Introduction

Currently, the epidemics of HIV-infection transferred to another stage – a stage of comorbid and severe forms of the disease. In these conditions, with the development of immunodeficiency an opportunistic infections of the lungs join in such as pulmonary tuberculosis and mycobacteriosis of the lungs, moniliasis, pneumocystic, bacterial and viral pneumonia accompanied with high lethality despite the use of antiretroviral treatment [3, 15].

Comorbidity of pulmonary tuberculosis and the HIV-infection is a classic model of simultaneous development of concomitant abnormality leading to serious complications and lethality [1, 5, 17]. It is determined by a high incidence of tuberculosis in population. So, according to the World Health Organization 10 million people had tuberculosis in 2019 (range: 9.0 – 11.1 mln) and the lethality from the disease was 1.2 mln (range: 1.2 – 1.3 mln) of people. Two hundred fifty one thousand lethal cases (range: 223 – 281 thous.) included HIV-patients, 85% of whom had not received any antiretroviral treatment, but the data on comorbidity with pulmonary tuberculosis and other opportunistic infections of the lungs as well as bacterial pneumonia are not given [24].

In the Russian Federation a part of first-time diagnosed patients with tuberculosis in 2010 was 6.1% and in 2020 it increased to 30.1%, of which the number of lethal cases from all causes changed from 11.7% to 14.2%, consequently. The situation was complicated with the increase of the part of the patients with tuberculosis at the late stages of HIV-infection with the amount of CD4+ lymphocytes less than 200 in 1 mcl of blood, it increased from 11.3% in 2010 to 25.6% in 2020, but there is no statistics on the comorbidity of tuberculosis and opportunistic infections of the lungs [13].

There are publications on clinical manifestations and diagnostics of bacterial pneumonia in patient with HIV-infections and various immune status, including the late stages with significant immunodeficiency (AIDS) in the foreign and Russian literature. In these cases colonization of the upper respiratory tract with various pathogenic bacteria leads to the development of bacterial pneumonia associated with the most common agents such as *Streptococcus pneumoniae* (*S. pneumoniae*) and *Staphylococcus aureus* (*S. aureus*) by endogenic way [2, 4, 20, 29].

The accent is put to the community-acquired pneumonia caused mainly by *S. pneumoniae* [5, 12, 14, 21, 23, 25, 27, 28, 30, 33, 34]. But in the international classification ICD-10 and the Russian Clinical Guidance on Community-acquired pneumonia in adults bacterial pneumonia in patients with HIV-infections and AIDS are put separately, since with significant immunodeficiency pneumonia associated with *S. aureus* may develop simultaneously, which is more consistent with hospital pneumonia [7, 18]. These documents and the new international classification ICD-11 have a classification of community-acquired and hospital pneumonia only in HIV-negative patients [19]. At the same time there is a complete lack of publications on the specifics of clinical manifestations and diagnostics of comorbidity of tuberculosis of the respiratory system, pneumonia associated with *S. pneumoniae* or *S. aureus* in patients with late stages of HIV-infection with immunodeficiency, which is an urgent issue for the studies of pulmonary tuberculosis and all clinical medicine in terms of etiological diagnostics and determination of drug resistance for prescription of timely treatment.

Current publication is a continuation of our priority combined studies on the clinics and diagnostics of comorbidity of pulmonary tuberculosis and opportunistic infections of the lungs in patients with HIV-infection and immunodeficiency [10, 11, 33]. We pay special attention to the lung damages caused immediately by HIV-infection manifesting in lymphoid and non-specific interstitial pneumonia. High incidence and advance of chronic obstructive lung disease (COLD), which in its case makes conditions for development of bacterial pneumonia [9, 26, 31]. Also, for the first time the complexity issues for etiological diagnostics of comorbid abnormality using the complex of modern laboratory and instrumental methods are being discussed as they have not been studied at all.

### Purpose

To study specifics of diagnostics and clinics of comorbidity of pulmonary tuberculosis and pneumonia associated with *S. pneumoniae* or *S. aureus* in patients with HIV-infection with immunodeficiency.

## Materials and methods

Ninety-three first-time diagnosed patients with pulmonary tuberculosis and 4B stage of HIV-infection in the advanced phase in the absence of antiretroviral therapy age 26-50 were included in the study. There were 61 male (65.6 ± 4.9%) and 32 females (34.4 ± 4.9%). The patients were divided into 3 groups. The 1^st^ group included 31 patient with pulmonary tuberculosis and pneumonia associated with *Streptococcus pneumoniae*, the 2^nd^ group included 31 patient with pulmonary tuberculosis and pneumonia associated with *Staphylococcus aureus*. The 3^rd^ group included 31 patient without bacterial pneumonia selected by a copy-pair principle. Patients in all groups were almost identical in terms of age, gender, social, clinical parameters, concomitant abnormality and stages of HIV-infection and immunodeficiency.

A mandatory criterium for the patients with pulmonary tuberculosis to be included in the study was culture isolation of *M. tuberculosis* in the diagnostic material from the respiratory system (sputum, bronchoalveolar lavage, biopsy material received at bronchoscopy during the punctures of intrathoracic lymph nodes) and other organs (blood, urine, faeces and punctures of peripheral lymph nodes). Culture of the diagnostic material was performed on the dense Lowenstein-Jensen medium and in the automated system BACTEC MGIT 960 with determination of drug resistance of the acquired culture to the antituberculosis drug products by the method of absolute concentrations [16].

For etiological diagnostics of bacterial pneumonia a culture of the diagnostic material from the respiratory tract on special nutrition media was performed. The acquired cultures were studied for drug resistance to wide spectrum antibacterial agents with disk-diffused method or method of serial dilutions [7].

For diagnostics of opportunistic infections of the lungs associated with *Candida albicans* 1 type (*C. albicans*), *Mycobacterium nontuberculosis* (*M. nontuberculosis*), *Pneumocystis jiroveci* (*P. jiroveci*), *Herpes simplex virus* (*HVS*) and *Cytomegalovirus hominis* (*CMVH*), viral, bacterial, immunological and molecular-genetic studies were used (polymerase chain reaction – PCR) of the diagnostic material from the respiratory tract [9, 20, 26, 31].

All patients received clinical laboratory immunological (determination of CD4+ lymphocyte count with the method of flow cytofluorimetry and viral load by the count of HIV RNA copies in blood) and radial study including CT of the chest, magnetic resonance imaging (MRI), ultrasound study of the internal organs.

Statistical treatment of the data was performed using Microsoft Office Excel 2019 with calculation of the mean parameter in the group and the standard error of the mean confidence interval (CI). Confidence criterium p was determined by Student’s table. Difference between mean arithmetic parameters were considered valid with p < 0.05.

## Results

HIV-infection in 93 patients was revealed 6-9 years ago at age of 20 to 41. At the time of diagnostics 77 (82.8 ± 3.9%) patients contracted the diseases via parenteral way. All of them were followed up at an AIDS centre, which they almost did not visit due to a social alienation and lack of compliance to the studies and the treatment, they did not work and they did not have families. All patients suffered from a drug abuse, consumed alcohol and smoked. All patients had concurrent abnormality – viral hepatitis B or C (in all patients) and COLD in 34 (36.6 ± 4.8%).

Pulmonary tuberculosis in 93 patients with HIV-infection was revealed during the referral because of the symptoms of acute inflammatory respiratory disease in the treatment facility of primary medical care of to the AIDS centre and was confirmed during a combined examination at an outpatient facility for tuberculosis treatment, where *M. tuberculosis* was isolated from the respiratory tract. Patients’ age by the time of pulmonary tuberculosis confirmation was 26-50 years old. From the time of HIV-infection confirmation to the tuberculosis confirmation number of patients with COLD increased to 52 (55.9 ± 5.0%) cases. All the patients were hospitalized to an inpatient unit for tuberculosis treatment to the special department for patients with tuberculosis with HIV-infection, where at admission *S. pneumoniae* or *S. aureus* were isolated from a diagnostic titre in the material from the respiratory tract.

Thus, comorbidity of pulmonary tuberculosis, bacterial pneumonia in patients with 4B stage of HIV-infection, in the advanced phase and with the absence of antiretroviral treatment, was diagnosed in 6-9 years after the confirmation of HIV-infection in the reproductive age in patients who did not work, did not have families, suffered from a drug abuse, consumed alcohol and smoked.

Patients’ distribution in the observed groups by CD4+ lymphocytes count per 1 mcl of blood is presented in **Table 1**.

**Table 1.** Patients’ distribution in the observed groups by CD4+ lymphocytes count per 1 mcl of blood.

| Groups | Number of patients | CD4+ lymphocytes count per 1 mcl of blood |  |  |  |
| --- | --- | --- | --- | --- | --- |
|  |  | 50-30 | 29-20 | 19-10 | <9 |
| 1 | Abs. 31<br>% 100 | 6<br>19.3±7.1 | 10<br>32.2±8.4 | 12<br>38.7±8.7 | 3<br>9.6±5.3 |
| 2 | Abs. 31<br>% 100 | 2<br>6.4±4.4 | 10<br>32.2±8.4 | 14<br>45.2±8.9 | 5<br>16.1±7.0 |
| 3 | Abs. 31<br>% 100 | 9<br>29.0±8.1 | 14<br>45.2±8.9 | 7<br>22.6±7.5 | 1<br>3.2±3.2 |

As shown in Table 1, in the observed group CD4+ lymphocyte count per 1 mcl of blood almost did not differ. In the 1^st^ group in 19.3% of patient CD4+ lymphocytes count was in the range of 50-30 cl./mcl of blood, in 32.2% – 20-29, in 38.7% – 19-10 and in 9.6% – less than 9, in the 2^nd^ group in 6.4%, in 32.2%, in 45.2% and in 16.1%, respectively, and in the 3^rd^ group – in 29% - 50-30 cl/mc, in 45.2% - 20-29, in 22.6% – 19-10 and in 3.2% - less than 9 (p > 0.05). Average count of CD4+ lymphocytes in 1 mcl of blood was also similar and was 28.3 ± 0.38 cl/mcl of blood in the 1^st^ group of patients, 24.7 ± 0.29 in the 2^nd^ group of patients and 31.7 ± 0.43 (p > 0.05) in the 3^rd^ group of patients. The viral load was more than 500 000 RNA HIV copies/ml of blood.

In patients of all groups pulmonary tuberculosis was characterized with generalization and multiple extrapulmonary damages, confirmed with isolation of M.tuberculosis from the diagnostic material from various organs. In the 1^st^ group two organs were damaged in 11 patients, three in 6 patients, four in 2 two patients and five in two patients; in the 2^nd^ and the 3^rd^ groups damage of two organs was in 10 and 12 patients, three in 8 and 5 patients, four in 1 and 2 patients and five in 1 and 2 patients, respectively (p>0.05).

Thus, patients with comorbidity of pulmonary tuberculosis and bacterial pneumonia and patients with pulmonary tuberculosis with 4B stage of HIV-infection in the advance phase with the absence of antiretroviral treatment had significant immunodeficiency with significant decrease of CD4+ lymphocyte count in blood (from 50 cells and less per 1 mcl of blood with its average count not exceeding 40 cl/mcl of blood), which is, to a significant degree, is associated with generalization of tuberculosis and determined similarity of clinical and radiological manifestations of comorbidity of pulmonary tuberculosis and bacterial pneumonia with isolated pulmonary tuberculosis.

At microbiological and PCR study of biological material from the respiratory tract other opportunistic infections of the lungs were also diagnosed confirmed by an isolation of a certain agent (**Table 2**).

**Table 2.** Patients’ distribution by groups according to the incidence of opportunistic infections of the lungs and lungs aetiology (M ± m).

| Groups | Number of patients | Opportunistic infections of the lungs: |  |  |  |  |
| --- | --- | --- | --- | --- | --- | --- |
|  |  | MP | PCP | MBL | HVP | CMVP |
| 1 | Abs. 31<br>% 100 | 15<br>48.4±8.9 | 7<br>22.6±7.5 | 7<br>22.6±7.5 | 9<br>29.0±8.1 | 4<br>12.9±5.9 |
| 2 | Abs. 31<br>% 100 | 19<br>63.3±8.6 | 8<br>25.8±7.6 | 6<br>19.3±7.1 | 10<br>32.2±8.4 | 3<br>9.7±5.3 |
| 3 | Abs. 31<br>% 100 | 15<br>48.4±8.9 | 8<br>25.8±7.6 | 9<br>29.0±8.1 | 8<br>25.8±7.6 | 6<br>19.3±7.1 |

*Note:* MP – moniliasis pneumonia, PCP – pneumocystic pneumonia, MBL – mycobacteriosis of the lungs, HVP – herpes virus pneumonia, CMVP – cytomegaloviral pneumonia.

As seen from **Table 2**, comorbidity with other opportunistic infections of the lungs was additionally revealed in patients in the observed groups, the incidence of which by the groups did not significantly differ: moniliasis pneumonia (MP), with isolation of *C. albicans*, was diagnosis in the 1^st^ group in 48.8% cases, in the 2^nd^ group in 63.3% and in the 3^rd^ group – in 48.8%; pneumocystic pneumonia (PCP) with isolation of *P. jiroveci* in 22.6%, 25.8% and 25.8%, respectively; mycobacteriosis of the lungs (MBL) with isolation of *M. nontuberculosis* (as a rule *M. Avium* complex) in 22.6%, 19.3% and 29%; herpes virus pneumonia (HVP), with isolation of HVS – in 29%, 32.2% and 25.8% and cytomegalovirus pneumonia (CMVP) with isolation of CMVH in 12.9%, in 9.7% and in 19.3% cases.

Consequently, in all patients both with comorbidity of pulmonary tuberculosis and bacterial pneumonia and pulmonary tuberculosis without bacterial pneumonia with 4B stage of HIV-infection with immunodeficiency in the advanced phase with the absence of antiretroviral treatment tuberculosis had generalized character with extrapulmonary damages of various organs and in some patients opportunistic infections of the lungs were diagnosed, which determines similarity of clinical manifestations and makes their differentiation difficult.

Clinical picture in patients of all groups almost did not differ and was characterized with significant syndrome of intoxication, fever, chills, weight loss, adynamic, headache, myalgia, tachycardia, pale skin. Respiratory symptoms in patients of all groups also did not differ significantly and were presented with dyspnoea, cough, excretion of mucous purulent sputum and presence of rales of various calibre in the lungs. In patients of 1^st^ and 2^nd^ groups if *S. pneumoniae* or *S. aureus* were isolated from the diagnostic material from the respiratory tract blood, the cough was more significant, sputum had purulent character, there was blood in the cough and increasing pulmonary cardiac failure. Laboratory studies showed parameters of inflammation characteristic for septic state: high leucocytosis >15 × 10^9^/l in the blood test, lymphopenia < 1.2 × 10^9^/l and abruptly increased ESR > 40 mm/h, and biochemistry showed high parameters of C-reactive protein < 100 mg/l.

Thus, in patients with 4B stage of HIV-infection with immunodeficiency in the advanced phase with the absence of antiretroviral treatment in combination with pulmonary tuberculosis and bacterial pneumonia or pulmonary tuberculosis without bacterial treatment the clinical picture was characterized with intoxication syndrome, respiratory manifestations, however, differences may be determined only with etiological diagnostics and isolation of certain agent of bacterial pneumonia and opportunistic infection of the lungs.

Patients’ distribution in the observed groups by the incidence and the character of drug resistance of *M. tuberculosis* to antituberculosis treatment in presented in **Table 3**.

**Table 3.**
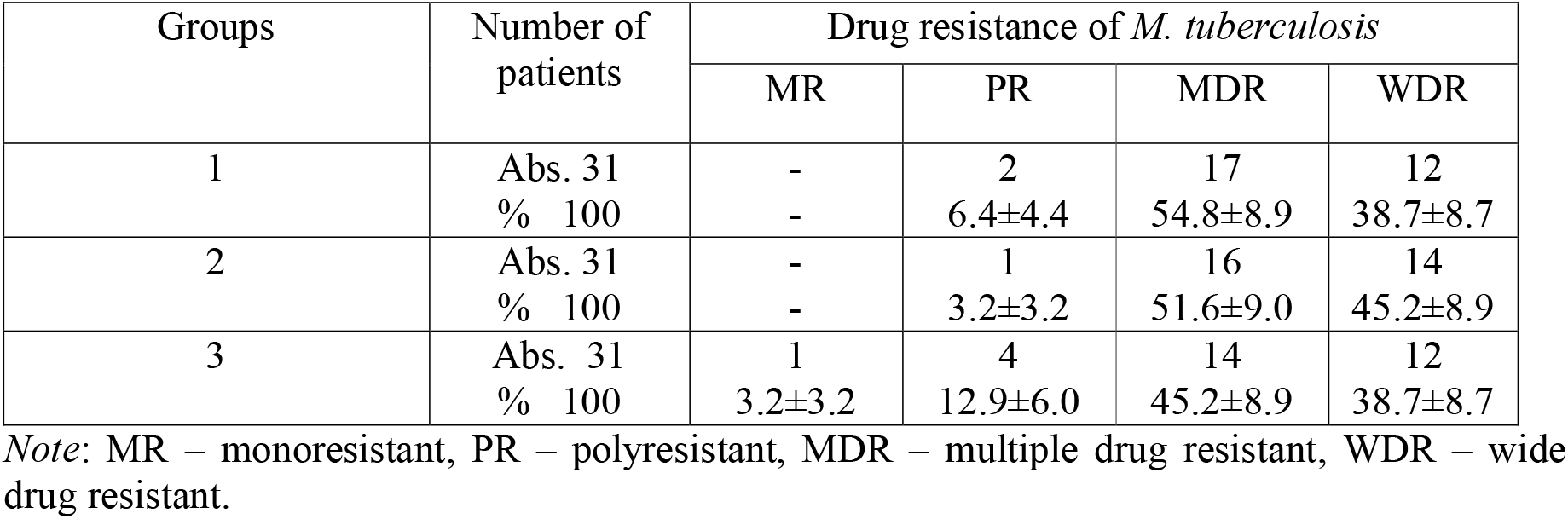
Patients’ distribution in groups by the incidence and the character of drug resistance of *M. tuberculosis* to antituberculosis treatment (M ± m).

| Groups | Number of patients | Drug resistance of *M. tuberculosis* |  |  |  |
| --- | --- | --- | --- | --- | --- |
|  |  | MR | PR | MDR | WDR |
| 1 | Abs. 31<br>% 100 | -<br>- | 2<br>$6.4 \pm 4.4$ | 17<br>$54.8 \pm 8.9$ | 12<br>$38.7 \pm 8.7$ |
| 2 | Abs. 31<br>% 100 | -<br>- | 1<br>$3.2 \pm 3.2$ | 16<br>$51.6 \pm 9.0$ | 14<br>$45.2 \pm 8.9$ |
| 3 | Abs. 31<br>% 100 | 1<br>$3.2 \pm 3.2$ | 4<br>$12.9 \pm 6.0$ | 14<br>$45.2 \pm 8.9$ | 12<br>$38.7 \pm 8.7$ |

Note: MR – monoresistant, PR – polyresistant, MDR – multiple drug resistant, WDR – wide drug resistant.

As seen from **Table 3** polyresistance was seen in the 1^st^ group in 6.4% of patients, in the 2^nd^ group in 3.2% and in the comparison group in 12.9% (p > 0.05), monoresistance (MR) was in only 1 patient (3.2%) in the comparison group, no *M. tuberculosis* sensitive to all antituberculosis treatment was found in the observed groups.

In most patients in the observed groups *M. tuberculosis* had multiple (MDR) and wide drug resistance (WDR). MDR in the 1^st^ group was revealed in 54.8% of cases, in the 2^nd^ group in 51.6% and in the 3^rd^ group in 45.2% (p > 0.05) and WDR, respectively, in 38.7%, 45.2% and 38.7% of cases (p > 0.05).

We had earlier described a high level of MDR *M. tuberculosis* in patients with caseous pneumonia with significant immunodeficiency when mean CD4+ lymphocyte count was less than 30 cl./mcl of blood [8]. Significant immunodeficiency in patients with HIV-infection was aggravated with extended caseous-necrotic lung damage determining severe clinical manifestations of the disease. Control of immune system over an unrestrained multiplication of *M. tuberculosis* was lost when incidence of spontaneous and induced mutations with formation of drug resistance abruptly increased. Such mutations occur in natural “wild” strains prior to contact with antituberculosis treatment and develop in the process of inadequate chemotherapy of tuberculosis with formation of MDR *M. tuberculosis* [8]. Such mechanism of development of MDR and WDR *M. tuberculosis*, is most likely realized with expressed immunodeficiency at the late stages of HIV-infection in patients with comorbidity of pulmonary tuberculosis and bacterial pneumonia.

At the same time in patients with caseous pneumonia with significant immunodeficiency pathogenic bacterial agents were isolated in the diagnostic titre from the respiratory tract with the culture method, including *S. pneumoniae* and *S. aureus*, with high level of resistance to wide spectrum antibacterial agents which significant aggravated the course of caseous pneumonia [8].

Patients in the 1^st^ group had multiple resistance of *S. pneumoniae* to beta-lactam antibacterial agents, tetracyclines, macrolides, levomycetin and other wide-spectrum antibacterial agents and to the drug products used for treatment of tuberculosis: rifampicin, levofloxacin, moxifloxacin, amoxicillin clavulanic acid, clarithromycin and meropenem, in patients in the 2^nd^ group – *S. Aureus* turned out to be multiple resistant to many wide spectrum antibacterial agents, in 8 (25.8%) of patients Methicillin-resistant (MRSA) and Vancomycin-resistant (VRSA) agents were revealed. Only linezolid turned out to be highly effective drug product in treatment of patients with bacterial pneumonia and tuberculosis associated with multiple resistant *S. pneumoniae*, MDR and WDR *M. tuberculosis*.

At the chest CT in patients of all three groups a combination of three abnormal syndromes [10, 11] was visualized. Syndrome of hematogenic and bronchogenic dissemination in the lungs with foci of various sizes (from small to large ones) and various intensity (from low to high), with tendency to fusion and formation of infiltrates with development of destruction of the lung tissue. Syndrome of intrathoracic lymphadenopathy with bilateral enlargement of intrathoracic lymph nodes with infiltrative changes along the periphery. Syndrome of the lung pattern change by the type of interstitial pneumonia is characterized with induration of interstitial tissue, diffuse decrease of lung transparency, diffused and increased lung pattern that has “reticular” character with areas of opaque glass.

At the same time in patient of the 1^st^ and the 2^nd^ groups infiltrates were visualized more frequently with bronchial lumen predominantly in the lower lobes of the lungs with formation of small abscesses, occurrence of exudate in the pleural cavity, however, these changes also occurred in patients in the 3^rd^ group. In these cases abnormalities associated with HIV-infection itself such as lymphoid interstitial pneumonia and non-specific interstitial pneumonia. Development of such changes connected with manifestations of opportunistic lung infections is not excluded: mycobacteriosis of the lungs, pneumocystic pneumonia, moniliasis pneumonia, herpes virus pneumonia and cytomegalovirus pneumonia. Differentiation of the nosologic aetiology of these changes of the chest organs was possible only with isolation of *M. tuberculosis, S. pneumoniae* and *S. aureus* or agents for other opportunistic lung infections.

## Discussion

Comorbidity of 4B stage of HIV-infection with immunodeficiency in the advanced phase with absence of antiretroviral treatment and pulmonary tuberculosis with isolation of *M. tuberculosis* combined with bacterial pneumonia associated with *S. pneumoniae* or *S. aureus*, or pulmonary tuberculosis without bacterial pneumonia is diagnosed in 6-9 years after the confirmation of HIV-infection in people of reproductive age, not working, not having families, suffering from a drug abuse, using alcohol and smoking with concurrent viral hepatitis B or C and COLD.

Comorbidity is characterized with expressed immunodeficiency (CD4+ lymphocyte count less than 50 cells per 1 mcl of blood and with mean parameter no more than 30 cl./mcl of blood), generalization of tuberculosis and presence of opportunistic infections of the lungs caused by *M. nontuberculosis, P. jiroveci, C. albicans, Herpes simplex virus* and *Cytomegalovirus hominis*. Clinical picture is characterized with a syndrome of inflammatory intoxication and respiratory manifestations. Chest CT reveals syndromes of lung dissemination, intrathoracic lymphadenopathy and changes of the lung pattern by the type of interstitial pneumonia. It is not possible to differentiate the nosologic aetiology of clinical and radiologic manifestations of the diseases.

Significant immunodeficiency in patients with 4B stage of HIV-infection with immunodeficiency in the advanced phase with the absence of antiretroviral treatment aggravated with severe course of caseous pneumonia or comorbidity of pulmonary tuberculosis and bacterial pneumonia determines a high degree of development of MDR and WDR *M. tuberculosis* to antituberculosis treatment and multiple resistance of *S. pneumoniae* and *S. aureus* to wide spectrum antibacterial treatment.

It should be said that Federal Clinical Guidance for Diagnostics and Treatment of tuberculosis [16] in patients with HIV-infection for treatment of patient with respiratory tuberculosis with isolation of MDR and WDR *M. tuberculosis* used IV and V modes of chemotherapy that include drugs being wide spectrum antibacterial agents and having activity against *M. tuberculosis*: kanamycin, amikacin, capreomycin, levofloxacin, moxifloxacin, linezolid, amoxicillin clavulanic acid, clarithromycin and meropenem, according to Clinical Recommendations of Russian Respiratory Society (RRS) and Interregional Association on Clinical Microbiology and Antimicrobial Chemotherapy (IACMAC) used for treatment of pneumonia associated with *S. pneumoniae* or *S. aureus* [7].

High incidence of drug resistance of *M. tuberculosis, S. pneumoniae* and *S. aureus* makes us doubt efficiency of empirical indication of the drug products included in IV and V modes of chemotherapy of tuberculosis and antibacterial treatment of bacterial pneumonia which requires determination of drug resistance to all drugs recommended for treatment of comorbidity of pulmonary tuberculosis and bacterial pneumonia in each case. Development of individual modes of chemotherapy of pulmonary tuberculosis and antibacterial treatment of bacterial pneumonia in patients with late stages of HIV-infection with immunodeficiency is recommended.

## Conclusion

Patients with HIV-infection specifically the late stages with significant immunodeficiency in case of pulmonary tuberculosis require microbiological and molecular genetic studies of diagnostic material from the respiratory tract in order to reveal the agents of bacterial pneumonia, including *S. pneumoniae* or *S. aureus*, and other opportunistic infections of the lungs and study of drug resistance of revealed agents considering the high incidence revealed in more than 80% of cases, MDR and WDR *M. tuberculosis* and multiple resistance in *S. pneumoniae* and *S. aureus* to many wide spectrum antibacterial agents for timely prescription of adequate etiological treatment and decrease of lethality in the category of such patients with comorbid disease.

## Data Availability

All data produced in the present work are contained in the manuscript

## Author Credentials

Mishina Anastasiya Vladimirovna - Holder of first-level doctoral degree (Kandidat nauk) in medicine, Associate Professor at the Department of Phthisiology and Pulmonology at the FSBEI HE The A.I. Yevdokimov Moscow State University of Medicine and Dentistry of the Ministry of Health of Russia, Moscow, Russia and the physician of the Department for patients with tuberculosis and HIV infection of FBHI The Tuberculosis Clinical Hospital N 3 named after Professor G.A. Zakharyin of the Ministry of Health of Russia, Moscow, Russia

Mishin Vladimir Yur’yevich - Holder of second-level doctoral degree (Doktor nauk) in medicine, Professor, Head of the Department of Phthisiology and Pulmonology at the FSBEI HE The A.I. Yevdokimov Moscow State University of Medicine and Dentistry of the Ministry of Health of Russia, Moscow, Russia and Consultant Professor at FBHI The Tuberculosis Clinical Hospital N 3 named after Professor G.A. Zakharyin of the Ministry of Health of Russia, Moscow, Russia

Shashenkov Ivan Vasil’yevich - Assistant Professor at the Department of Phthisiology and Pulmonology of FSBEI HE The A.I. Yevdokimov Moscow State University of Medicine and Dentistry of the Ministry of Health of Russia, Moscow, Russia

## References

1. Azovtseva O.V., Gritsyuk A.V., Gemaeva M.D., Karpov A.V., Arkhipov G.S. HIV infection and tuberculosis as the most complex variant of comorbidity. Bulletin of Novogorodsky State University. 2020, vol. 117, nn. 1, pp. 79–84. (In Russ.) DOI: https://doi.org/10.34680/2076-8052.2020.1(117).79-84

2. Akusheva D.N., Khokhlova O.E., Kamshilova V.V., Motova A.I., Peryanova O.V., Upirova A.A., Potkina N.K., Yamamoto T. Community-acquired pneumonia in HIV-infected people: microflora, antibiotic resistance, role in development depending on the level of CD4 lymphocytes. // Medical Immunology. - 2019. - Volume 21. - No. 3. - pp. 457-466. (In Russ.) DOI: 10.15789/1563-0625-2019-3-457-466

3. Belyakov N.A., Rassokhin V.V., Trofimova T.N., Stepanova E.V., Panteleev A.M., Leonov O.N., Buzunova S.A., Konovalova N.V., Milichkina A.M., Totolyan A.A. Comorbid and severe forms of HIV infection in Russia. HIV infection and immunosuppression. 2016, vol. 8, nn. 3. pp. 9-25. (In Russ.) DOI: https://doi.org/10.22328/2077-9828-2016-8-3-9-25

4. Bozoyan A.A., Puzyreva L.V. Features of bacterial pneumonia in HIV-infected people. // Crimean Therapeutic Journal. 2019, nn. 2, pp. 28-32. (In Russ.)

5. Borodulina E.A., Vdoushkina E.S., Borodulin B.E., Povalyaeva L.V. HIV infection and community-acquired pneumonia. Causes of death. HIV infection and immunosuppression. 2019, vol. 11, no. 1, pp. 56-63. (In Russ.) DOI: https://doi.org/10.22328/2077-9828-2019-11-1-56-63

6. Viktorova I.B., Zimina V.N., Dadyka I.V., Andreeva I.V., Golovina I.A., Chuzhikova E.P. Community-acquired pneumonia in patients with HIV infection. Tuberculosis and lung disease. 2021, vol. 99. nn. 4. pp. 22-28. (In Russ.) DOI: https://doi.org/10.21292/2075-1230-2021-99-4-22-28

7. Community-acquired pneumonia “MKB 10”: Clinical recommendations of the Russian Respiratory Society (RRS) and the Interregional Association for Clinical Microbiology and Antimicrobial Chemotherapy (IACMAC). 2018, 88 p. (In Russ.)

8. Erokhin V.V., Mishin V.Yu., Chukanov V.I., Giller D.B. Caseous pneumonia. M.: “Medicine”. 2008, 191 p. (In Russ.) Clinical recommendations. HIV infection in adults. National Association of Specialists in the Prevention, Diagnosis and Treatment of HIV Infection (approved. Ministry of Health of the Russian Federation). 2020, 230 p. (In Russ.)

9. Mishina A.V., Mishin V.Yu., Sobkin A.L., Osadchaya O.A. Disseminated and generalized pulmonary TB and opportunistic diseases in patients with late-stage HIV infection and immunosuppression. Tuberculosis and Lung Diseases, 2018, vol. 96, no. 12, pp. 68-70. (In Russ.) DOI: https://doi.org/10.21292/2075-1230-2018-96-12-68-70

10. Mishina, A. V., Mishin V. Yu., Ergeshov A. E., Sobkin A. L., Romanov V. V., Kononets A.S. clinical manifestations and diagnosis of a combination of tuberculosis of respiratory organs and opportunistic lung infections in adult patients in the later stages of HIV infection with immunodeficiency. CONSILIUM Medium. 2020? – vol. 22, nn. 11. pp. 78-86. (In Russ.) DOI: https://doi.org/10.442120751753.2020.11.200184

11. Puzyreva L.V., Mordyk A.V., Ovsyannikov N.V. Bacterial pneumonia in HIV-infected patients. Epidemiology and infectious diseases. Current issues. 2019 nn. 3, pp. 92-98. (In Russ.) DOI: https://dx.doi.org/10.18565/epidem.2019.9.3.92-8

12. Resources and activities of anti-tuberculosis organizations of the Russian Federation in 2019-2020 (statistical materials). / O.B. Nechaeva, I.M. Son, A.V. Gordina, S.A. Sterlikov, D.A. Kucheryavaya, A.V. Dergachev, S.B. Ponamarev M.: RIO TSNIIOIZ. 2021, 112 p. (In Russ.)

13. Sabitova R.Ya., Zhestkov A.V., Alpatova T.A. Community-acquired pneumonia in drug users with HIV infection: the peculiarities of clinical and laboratory manifestations. Atmosphere. Pulmonology and allergology, 2012, no. 3, pp. 23-27. (In Russ.)

14. Fazylov V.H., Manapova E.R., Akifyev V.O. Clinical and epidemiological characteristics of secondary diseases in HIV-infected patients in the conditions of inpatient care. // Infectious diseases: news, opinions, training. 2020, vol. 9, nn. 4, pp. 81-87. (In Russ.) DOI: https://doi.org/10.33029/2305-3496-2020-9-4-81-87

15. Federal clinical recommendations on diagnosis and management of TB in HIV-infected patients. Moscow-Tver, Triada, 2014, 56 p. (In Russ.)

16. Shuvalova E.V., Vishnevsky A.A. Comorbidity in patients with HIV infection and tuberculous spondylitis as a risk factor for infectious complications. Spine surgery. 2020, vol. 17. nn 1, pp. 96–101. (In Russ.) DOI: https://dx.doi.org/10.14531/ss2020.1.96-101

17. The electronic reference book of the international classification of diseases, the 10th revision (ICD-10), 1990-2020. Diagnosis codes: search by code, name, time :of emergence or form of disease. (In Russ.)

18. Electronic handbook of the International Classification of Diseases of the 11th revision (ICD-11) 2021. ICD classes 11. (In Russ.)

19. Bartlett G., Redfield R., Pham P. Bartlett’s Medical Management of HIV Infection. Oxford Univercity Press, 2019, 864 p. DOI: 10.1093/med/9780190924775.001.0001

20. Cilloniz C., Torres A., Polverino E., Gabarrus A., Amaro R., Moreno E., Villegas S., Ortega M., Mensa J., Marcos M.A., Moreno A., Miro J. Community-acquired lung respiratory infections in HIV-infected patients: microbial aetiology and outcome. European Respiratory Journal. 2014, vol. 43, nn. 6, pp. 1698–1708. DOI: 10.1183/09031936.00155813

21. Feinstein A. The pre-therapeutic classification of comorbidity in chronic disease. Journal of Chronic Diseases. 1970, vol. 23, no. 3, pp. 455–468.

22. Figueiredo-Mello C., Naucler P., Negra M., Levin A. Prospective etiological investigation of community-acquired pulmonary infections in hospitalized people living with HIV. Medicine (Baltimore). 2017, vol. 96, nn. 4, pp. 57–78. DOI: 10.1097/MD.0000000000005778

23. Global tuberculosis report WHO 2020. WHO. 2020, 232 p.

24. Godet C., Beraud G., Cadranel J. Bacterial pneumonia in HIV-infected patients (excluding mycobacterial infection). Revue des Maladies Respiratories. 2012, vol. 29, nn. 8, pp. 1058–1066.

25. Guidelines for Prevention and Treatment of Opportunistic Infections in HIV-Infected Adults and Adolescents with HIV. Recommendations from the Centers for Disease Control and Prevention,the National Institutes of Health, and the HIV Medicine Association of the Infectious Diseases Society of America. 2021, 479 p.

26. Horo K., Koné A., Koffi M-O, Ahui J., Brou-Godé C., Kouassi A., N’Gom A., Koffi N., Aka-Danguy E. Comparative Diagnosis of Bacterial Pneumonia and Pulmonary Tuberculosis in HIV Positive Patients. Rev. Mal. Respir. 2016, vol. 33, nn. 1, pp. 47–55. DOI: 10.1016/j.rmr.2015.01.004

27. Madeddu G., Fiori L, Mura S. Bacterial community-acquired pneumonia in HIV-infected patients. Curr. Opin. Pulm. Med. 2010, vol. 16, nn. 3. pp. 201–207. DOI: 10.1097/MCP.0b013e3283375825

28. Mendelson F., Griesel R., Tiffin N., Rangaka M., Boulle A., Mendelson M., Maartens G. C-reactive protein and procalcitonin to discriminate between tuberculosis, Pneumocystis jirovecii pneumonia, and bacterial pneumonia in HIV-infected inpatients meeting WHO criteria for seriously ill: a prospective cohort study. BioMed Central, 2018, nn.18, pp. 388–399. DOI: 10.1186/s12879-018-3303-6

29. Önür S. T., Dalar L., Sinem I., Yalçın A. Pneumonia in HIV-Infected Patients Eurasian J. Pulmonol. 2016, nn. 18. pp. 11–17. DOI: 10.5152/EJP.2016.81894

30. Panel on Opportunistic Infections in HIV-Infected Adults and Adolescents. Guidelines for the prevention and treatment of opportunistic infections in HIV-infected adults and adolescents: recommendations from the Centers for Disease Control and Prevention, the National Institutes of Health, and the HIV Medicine Association of the Infectious Diseases Society of America. Chapter «Bacterial Respiratory Disease». 2018, pp. 97–109.

31. Parekh A. M. B. Barton M. The challenge of multiple comorbidities for the US health care system. The Journal of the American Medical Association. 2010, – vol. 303, nn. 13, pp. 1303–1304. DOI: 10.1001/jama.2010.381

32. Peck K., Kim T., Min A., Kyung S., Han J. Pneumonia in immunocompromised patients: updates in clinical and imaging features. Precision and Future Medicine. 2018, vol. 2, nn. 3, pp. 95–108. DOI: 10.23838/PFM.2018.00121

33. Segal L., Methé B., Nolan A., Hoshino Y., Rom W., Dawson R, Bateman E, Weiden M. HIV-1 and bacterial pneumonia in the era of antiretroviral therapy. Proc. Am. Thorac. Soc. 2011, vol. 8, nn. 3, pp. 282–287. DOI: 10.1513/pats.201006-044WR

34. Mishina A.V., Mishin V.Y., Shashenkov I.V. CLINICAL MANIFESTATIONS AND DIAGNOSIS OF CO-INFECTION OF COVID-19, TUBERCULOSIS AND OPPORTUNISTIC PULMONARY INFECTIONS IN LATE-STAGE HIV PATIENTS WITH IMMUNODEFICIENCY medRxiv 2022.04.26.22274235; doi: https://doi.org/10.1101/2022.04.26.22274235

